# Triage of general practitioner referrals to internal medicine: identifying unnecessary referrals and exploring underlying referral reasons

**DOI:** 10.64898/2026.05.06.26352528

**Authors:** R.M.C. Pepping, R.C. Vos, H.M.M. Vos, M. E. Numans, M.O. van Aken

## Abstract

**Introduction:** Access to specialist care in the Netherlands requires a general practitioner (GP) referral, yet referrals to secondary care keep rising. Triage has been proposed to manage this demand and may be relevant for internal medicine, which addresses diverse and increasingly complex conditions. This study aimed to identify the internal medicine healthcare needs which were redirected to the GP after triage and to explore the factors driving GP referral behaviour.

**Methods:** This multi-method study combined quantitative referral data with qualitative insights from GP focus groups. Data were extracted from a hospital in an urban region, including adults with non-acute complaints referred for outpatient consultation to internal medicine between August 2019 and July 2021. Referrals were triaged for appropriateness and redirected where possible. Focus groups explored GPs’ perspectives on referral practices.

**Results:** Of 5,826 referrals triaged, 998 (17%) were redirected to the GP with advice and guidance. Endocrinology accounted for 35% of redirected cases, followed by nephrology (8.6%). Focus groups revealed underlying drivers of referral behaviour, identifying four themes: medical factors; GP-related factors, including professional uncertainty and autonomy; patient-related factors; and external factors, such as contextual and regulatory influences.

**Conclusion:** This study demonstrates that triage is a feasible strategy for managing referral volumes, particularly within domains such as endocrinology where many medical problems can be managed in primary care. However, referrals are shaped by more than clinical need, reflecting uncertainty, emotional considerations, patient expectations and systemic factors. Strengthened collaboration between primary and secondary care, alongside pre-referral consultation strategies, is essential to ensure appropriate, high-quality patient care.

## Introduction

Primary care is accessible for all residents of the Netherlands. The vast majority of the population is registered with a general practitioner (GP), often maintaining this relationship for decades. GPs typically serve as the primary healthcare provider and are the first point of contact for a broad range of healthcare needs, concerns and issues. They play a crucial role in coordinating care and are responsible for supporting optimal use of more expensive secondary care. Therefore, to access secondary specialist care, patients require a referral from their GP. In 2023, 78% of patients registered with a GP had at least one consultation, and 97% of these initial consultations were resolved without referral (1). Nonetheless, referrals to and use of secondary care has been rising since 2016 (1). This trend has triggered concern and research, particularly into the wide variation in referral rates amongst GPs. Previous studies have linked this variation to multiple factors, including patient characteristics, practice characteristics and gender of the GP (2–6).

To manage healthcare demand in secondary care, triage of GP referrals has been proposed as a form of pre-referral consultation (7). This approach could be particularly relevant for internal medicine, a broad specialty addressing many increasingly complex medical problems in an aging population. A recent study conducted in an urban region showed that approximately one in six (17%) triaged referrals to internal medicine could be managed without the need for a face-to-face consultation (8). In these cases, specialists primarily provided written advice on treatment and management in general practice. However, relatively little is known about the exact healthcare needs or domains that are redirected back to the GP after triage.

The Dutch Society of GPs, in collaboration with its partners, is responsible for the development and maintenance of national GP guidelines, over 100 of which concern treatment in primary care (9). Most of these guidelines explicitly outline the management options available within primary care and define appropriate referral. However, although referral guidelines describe when referral is appropriate, variation in referral decisions persists, suggesting that other factors play an important role. Understanding underlying drivers for this variation is essential, particularly in the context of triage systems that redirect referrals back to primary care.

To better understand referral practices and internal medicine healthcare needs, the present study addresses the following two objectives. First, it seeks to identify the initial healthcare needs referred to the department of internal medicine that are subsequently redirected to the GP. Second, it aims to explore the underlying drivers behind GP referrals, with a focus on those prone to redirection.

## Methods

### Study design and setting

This study used a multi-method approach to address both research questions. It builds on a previously published analysis of quantitative redirection data from the same hospital (8). For the present study, we supplemented these data with newly conducted focus groups with participating GPs, enabling a more comprehensive exploration of the factors underlying referral behaviour. The quantitative component drew on the internal medicine subset of the earlier hospital-wide mixed-methods study, in which referrals were triaged by a medical specialist and redirected back to the GP, with advice and guidance, when deemed appropriate (8). Triaging was performed by multiple dedicated internal medicine specialists who incorporated it into their work routine. All incoming referrals were assessed, especially regarding the expected added value of the secondary. When a referral to secondary care was considered unnecessary, a letter was sent and the medical specialist contacted the referring GP by telephone within 72 hours to explain the reasoning behind the decision to redirect and to offer advice. The qualitative component of the current study explored GPs’ referral practices and experiences using focus group discussions. Referral data were gathered from a hospital in an urban region of the Netherlands, serving a population of over 500,000 patients. Adults with non-acute health complaints referred for an outpatient consultation to the internal medicine department between August 2019 and July 2021 were included in this study.

### Data collection

Routine care data were pseudonymised and used to determine reasons for referral. Incoming referrals between August 2019 and July 2021 were extracted from electronic health records (EHR). GPs indicate a prespecified condition/complaint in the referral system. Data were collected on referral reason, patient characteristics, and whether an additional referral for the same complaint was made within the six-month follow-up period.

#### Focus groups with general practitioners

In total, 50 GPs from the hospital region were approached, of whom 13 (26%) agreed to participate. Ultimately, 11 participated, assigned to three focus groups. RP facilitated the focus groups, while CM took field notes. The focus groups were held in January and February 2025. The first group was joined by three GPs, the second and third by four. The third focus group was the only one held online. Participants all knew each other superficially, mainly through work, but were not direct colleagues, which enhanced speaking freely. Focus groups started with an introductory presentation of the quantitative results and earlier research (8), which was followed by the question: ‘What comes to mind when you consider referral, and what factors influence your decision to refer?’ Participants wrote their thoughts on sticky notes, grouped by RP into themes based on similarity. After no new sticky notes were added, these themes formed the basis for group discussion. Having discussed the general themes, the focus group was asked to consider two frequently cited referral reasons identified in the quantitative data. This part focused on referrals related to endocrinology, especially thyroid disorders, as well as nephrology, with a particular emphasis on decision-making related to referrals. National GP guidelines for thyroid disorders and renal insufficiency/chronic kidney disease were printed and distributed (9). GPs were asked to read through the guidelines and identify any difficult sections, or sections they usually refer to or deviate from, before discussing these with the group. The focus groups were structured the same each time. After three focus groups no new information was written on sticky notes and no new topics were mentioned during the discussion.

### Data analysis

The collected EHR data were analysed using SPSS version 28.0.1.0. Frequencies and descriptive statistics were calculated. Audio recordings of the focus groups were transcribed verbatim, and then coded and analysed with ATLAS.ti version 24.2.0.32043, using the steps recommended by Braune & Clark (10). First, all transcripts were read to allow the researchers to familiarize themselves with the data (phase 1). RP and CM coded all focus groups and MvA acted as third coder. Initial codes were generated inductively by RP (phase 2). The different codes were sorted into potential themes and code groups by RP and CM (phase 3). Data extracts within each theme were reviewed and refined, including merging and breaking down potential themes (phase 4). Clear definitions and final themes were then generated (phase 5). To minimize the risk of bias, during phases 4 and 5 data were discussed within the research group (RP, CM, MvA, HV) until consensus was reached on themes and definitions.

### Ethical considerations

The study was approved by the Medical Ethics Committee of Leiden Den Haag Delft (reference N21.093). The protocol was subsequently presented to the hospital ethics committee, who approved the protocol (reference T21-077). All interviewees gave informed consent to participate in the study. All methods were performed in accordance with the Declaration of Helsinki.

## Results

### Identifying internal medicine referrals

A total of 5,826 referrals to the department of internal medicine were assessed during triage. Of these, 998 (17%) were redirected to the GP with additional advice and treatment guidance (Table 1). Within a six-month follow-up period, only 86 (8.6%) patients were re-referred to hospital for the same care need. We found no difference in redirection rates between referrals of male and female patients (Table 2).

**Table 1.**
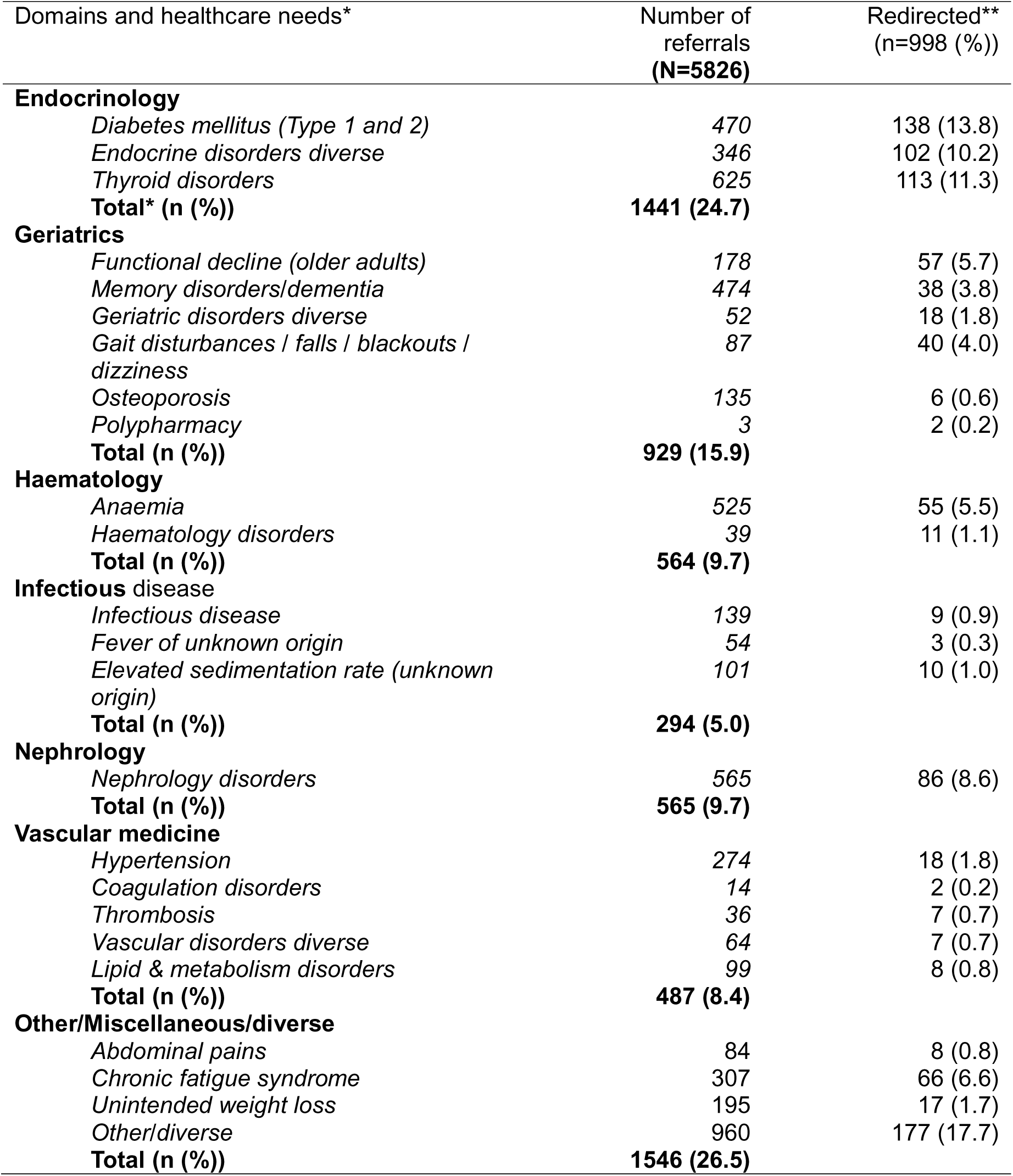
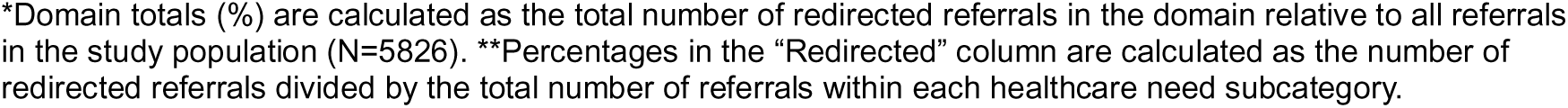
Referral and redirection rates per healthcare need and per internal medicine domain

| Domains and healthcare needs* | Number of referrals<br>(N=5826) | Redirected**<br>(n=998 (%)) |
| --- | --- | --- |
| **Endocrinology** |  |  |
| Diabetes mellitus (Type 1 and 2) | 470 | 138 (13.8) |
| Endocrine disorders diverse | 346 | 102 (10.2) |
| Thyroid disorders | 625 | 113 (11.3) |
| **Total\* (n (%))** | **1441 (24.7)** |  |
| **Geriatrics** |  |  |
| Functional decline (older adults) | 178 | 57 (5.7) |
| Memory disorders/dementia | 474 | 38 (3.8) |
| Geriatric disorders diverse | 52 | 18 (1.8) |
| Gait disturbances / falls / blackouts / dizziness | 87 | 40 (4.0) |
| Osteoporosis | 135 | 6 (0.6) |
| Polypharmacy | 3 | 2 (0.2) |
| **Total (n (%))** | **929 (15.9)** |  |
| **Haematology** |  |  |
| Anaemia | 525 | 55 (5.5) |
| Haematology disorders | 39 | 11 (1.1) |
| **Total (n (%))** | **564 (9.7)** |  |
| **Infectious disease** |  |  |
| Infectious disease | 139 | 9 (0.9) |
| Fever of unknown origin | 54 | 3 (0.3) |
| Elevated sedimentation rate (unknown origin) | 101 | 10 (1.0) |
| **Total (n (%))** | **294 (5.0)** |  |
| **Nephrology** |  |  |
| Nephrology disorders | 565 | 86 (8.6) |
| **Total (n (%))** | **565 (9.7)** |  |
| **Vascular medicine** |  |  |
| Hypertension | 274 | 18 (1.8) |
| Coagulation disorders | 14 | 2 (0.2) |
| Thrombosis | 36 | 7 (0.7) |
| Vascular disorders diverse | 64 | 7 (0.7) |
| Lipid & metabolism disorders | 99 | 8 (0.8) |
| **Total (n (%))** | **487 (8.4)** |  |
| **Other/Miscellaneous/diverse** |  |  |
| Abdominal pains | 84 | 8 (0.8) |
| Chronic fatigue syndrome | 307 | 66 (6.6) |
| Unintended weight loss | 195 | 17 (1.7) |
| Other/diverse | 960 | 177 (17.7) |
| **Total (n (%))** | **1546 (26.5)** |  |

\*Domain totals (%) are calculated as the total number of redirected referrals in the domain relative to all referrals in the study population (N=5826). \*\*Percentages in the "Redirected" column are calculated as the number of redirected referrals divided by the total number of referrals within each healthcare need subcategory.

**Table 2.** Distribution of redirected referrals

| Sex | Total<br>(N=5826) | Redirected (n/N, (%)) |
| --- | --- | --- |
| Male | 2141 | 349/2141 (16.3) |
| Female | 3685 | 649/3685 (17.6) |

The domain endocrinology was by far the largest, with 35% of referrals resulting in redirection. One-third involved thyroid-related concerns and another third concerned type 1 and 2 diabetes mellitus treatment. Thyroid gland referrals were diverse, often involving misinterpreted laboratory values (Table 3), many of which could have been resolved by adhering to primary care guidelines. Regarding nephrology referrals, 8.6% were redirected, distributed across albuminuria with impaired renal function and acute renal function decline (Table 4). Almost all were solved with advice and treatment guidance in general practice, without requiring a face-to-face consultation. The domain ‘other/miscellaneous/diverse’ covered a relatively large number of referrals. These consisted, for example, of healthcare needs related to multiple domains, a patient’s wish for a second opinion or a condition covered by a specified domain but where the GP chose the option ‘other’. Additionally, many referrals concerned unintended weight loss, chronic fatigue syndrome, abdominal pains and osteoporosis.

**Table 3.** Thyroid gland-related redirected referrals

| Thyroid disorder | Redirected referrals<br>(N=113, (n (%)) |
| --- | --- |
| Thyroid node | 37 (32.7) |
| Hypothyroidism | 31 (27.4) |
| Hyperthyroidism | 14 (12.4) |
| Subclinical hypothyroidism | 12 (10.6) |
| Subclinical hyperthyroidism/<br>postpartum thyroiditis | 11 (9.7) |
| Post (hemi) thyroidectomy | 5 (4.4) |
| Hyperparathyroidism | 2 (1.7) |
| Unknown | 1 (0.9) |

**Table 4.** Nephrology-related redirected referrals

| Nephrology disorder | Redirected referrals<br>(N=86, (n (%)) |
| --- | --- |
| Albuminuria + ACR > 30 | 27 (31.4) |
| Albuminuria + ACR 3-30 and eGFR<br>>30 | 25 (29.0) |
| (suspected) Acute kidney injury | 18 (20.9) |
| Deterioration of kidney function in CKD | 13 (12.1) |
| Albuminuria ACR 3-30 and DMt2 | 2 (2.3) |
| Type 1 Renal tubular acidosis | 1 (1.2) |

ACR: albumin-to-creatinine ratio (mg/mmol), eGFR: estimated glomerular filtration rate (ml/min/1.73 m<sup>2</sup>), CKD: chronic kidney disease, DMt2: Diabetes mellitus type 2.

### Qualitative focus groups analysis

In-depth focus groups were conducted to explore underlying reasons for referral. Between January and February 2025, three focus groups involving a total of 11 GPs were held. The mean age was 44.6 years, with six males and five females. Participants had on average 14 years’ experience as GPs, ranging from 1 to 40 years. Six were practice owners. The focus groups were structured the same way. Discussions about the apparent themes were lively and very interactive, even with the relative low number of participants.

Our focus groups revealed various drivers for referral, which were categorized into four main themes, each representing determinants influencing the referral decision. Two themes were further subdivided into subgroups. The identified themes were: 1) medical drivers, 2) general practitioner-related drivers, subdivided into ‘professional uncertainty’ and ‘autonomy’, 3) patient-related drivers and, 4) external drivers, subdivided into ‘contextual’ and ‘regulatory’ factors. While some factors were closely related, GPs assigned distinct meanings to them. The four themes were grouped into two overarching domains: medical and non-medical.

#### Theme 1. Medical drivers

Focus group participants largely agreed on medical drivers of referral. Some even failed to explicitly record these on sticky notes, viewing them as self-evident. These drivers reflected routine medical considerations and aligned closely with clinical guidelines. This topic generated little debate, although GPs differed concerning preferences and ‘comfortable’ domains. For example, some felt comfortable managing hypothyroidism during pregnancy, while others referred promptly. Several GPs also mentioned that they referred older adults or complex patients with multimorbidity earlier than guidelines suggest, yet still considered this a medical decision. Other commonly-cited drivers included the need for advanced diagnostic evaluation, exclusion of pathology, or access to specialist expertise in secondary care. ‘Gut feeling’ or ‘clinical intuition’ was also regarded as a medical driver. Participants noted a subtle but relevant distinction between intuitive clinical judgment and the formal referral indications for further investigation or specialist care to exclude serious pathology.

> FG1: I have the impression that the number of indications for referral is increasing and that requirements are becoming ever stricter?
>
> FG3: If I find, say, clear laboratory abnormalities that require further diagnosis or if I just feel an abdominal lump or an enlarged liver, then I refer.

#### Theme 2. General practitioner-related drivers

This theme was broad and was therefore divided into two subgroups: *professional uncertainty* and a*utonomy*.

<u>Professional uncertainty</u> included drivers such as doubts concerning the treatment initiated in general practice - particularly when no improvement or progress is evident. Even when a treatment resulted in improvement or progress, some GPs still sought additional reassurance regarding treatment. During discussions, participants emphasized that seeking confirmation and uncertainty due to lack of experience were important drivers of referral. A commonly-mentioned example regarding *insufficient experience* was hypothyroidism during pregnancy. Although GPs may treat such cases themselves, many prefer to refer due to limited experience or lack of confidence.

> FG2: Sometimes you see someone you’ve started on medication whose symptoms persist, which can, of course, be a reason to refer them. Self-reassurance is also a factor.
>
> FG1: As concerns pregnancy and thyroid problems - we don’t see these very often. I don’t know if I’d treat them myself. Maybe, but I would definitely need to consult the guidelines.

<u>Autonomy</u> includes drivers that are more personal in nature. GPs stressed the importance of maintaining their autonomy in the referral process; the decision to refer is theirs. This sense of autonomy also carried emotional weight. As independent professionals they expect their decisions, including referrals, to be taken seriously. In the context of doctor–patient relationships, participating GPs stated that they sometime refer not because they support the referral itself, but because it helps to maintain or restore the relationship. In such cases, the referral enables them to continue care in the primary care setting after referring. This differs from patient-driven referrals, discussed under patient-related drivers, although both may result in a referral being necessary to proceed effectively in primary care.

> FG3: You also feel a little criticized concerning your professionalism when you receive comments about a referral. I sometimes know that it is not strictly necessary, but I still decide to refer.

#### Theme 3. Patient-related drivers

One of the most important patient-related drivers for referral is the patient’s explicit demand to be referred to secondary care. Almost all participants noted this. This demand seems linked to an attitude of entitlement regarding access to hospital-based care. In some cases, these demands could be managed with thorough explanation and reassurance. But at times, when patients keep insisting it becomes increasingly difficult to resist referring, even in the absence of a medical rationale. These situations sometimes overlap with ‘professional uncertainty’, as persistent demands caused some GPs to question their own judgement, *‘Am I missing something? What if there is some pathology?’* Such demanded referrals are typically not clinically necessary but are designed to avoid conflict. When explanation fails, some GPs may concede. However, some GPs noted that not all patients are satisfied after a referral — some may remain dissatisfied and request another referral to a different hospital or specialist. In such cases, GPs sometimes noted in the referral system that the referral is made “at patient’s request”, signalling to the specialist that they did not support the referral, but saw no alternative in order to move forward in the doctor-patient relationship. In addition, cultural factors also influenced referral behaviour, particularly in situations where patients’ expectations of healthcare differ from the reality of the Dutch healthcare system.

> FG1: It’s definitely also about people becoming increasingly demanding. Yes, people are becoming more insistent. They say, "Yes, I have a regular headache but I want to see that particular migraine doctor, the neurologist; that’s who I want to see."
>
> FG2: But then, then they come three times for their thyroid. And eventually they want to see the endocrinologist, because, well, he’s so much better. And then, and that’s the worst of it, they want to see another endocrinologist because the first one couldn’t do anything either and he didn’t even listen.
>
> FG3: I have a few patients (who say), “I want to see the orthopaedist for my shoulder”, and I think, "God, I’ll have to see them six more times (if I don’t agree). ". That’s it, that means I’m actually more likely to refer. Because you know that the patient will otherwise just keep insisting. You know how it’s going to play out.

#### Theme 4. External drivers

This theme includes a range of aspects, categorized into two subgroups: *contextual* and *regulatory drivers*.

<u>Contextual</u> drivers relate to how care is organised. GPs reported that increasingly short outpatient trajectories and limited continuity in specialist care sometimes leads to repeated or premature referrals. For instance, when patients are discharged after a brief consultation or hospital admission, follow-up often reverts to the GP, who may feel the need to re-refer when problems persist. Similarly, the fragmentation of care, particularly when managing complex or chronic conditions, was mentioned as a challenge that can trigger additional referrals, even when medical necessity is unclear. Furthermore, a rising workload and time constraints were also cited as drivers influencing referral behaviour. The growing intensity and pace of general practice sometimes left insufficient time for extended diagnostics or complex decision-making, increasing the likelihood of referring to secondary care as a practical solution. In addition, although not a driver in itself of the decision to make a referral, the actual time required to write the referral request is taken into account. Some participating GPs indicated that they prefer to write a good, complete and concise referral, but that this takes time. Similarly, the referral system is not always supportive of the most efficient or convenient approach. Missing clinical information or limited upload options can prolong the process, which demands even more time.

> FG1: because more patients have ended up in primary care, you’re ultimately re-referring more patients? People are admitted for shorter periods and some things have to be re-referred, etc..
>
> FG2: I remember the time when as a GP you’d just look at a throat, a knee, or someone’s abdomen. Nowadays, it sometimes feels like we’re expected to manage all hospital care that doesn’t require an operating room, so to speak. And then I think, okay, you know, you really are expecting too much of me.

<u>Regulatory</u> drivers involve formal requirements imposed by national policies and regional guidelines and agreements. In some instances, referrals were initiated not because the GP felt it was needed, but because protocols or healthcare insurers require specialist documentation for treatment or reimbursement. Some referrals are ‘defensive’, driven by legal concerns or the need to share responsibility. In this context, adhering to rules or avoiding liability could become a dominant reason for referral, even when the GP would prefer to manage the case within primary practice. For some this linked to professional uncertainty, while others stated this felt different due to the origin of the referral.

> FG1: You know, from the moment of referral, when there is a long waiting list, I would really like to place the responsibility with the secondary care provider. Legally, I know that’s not the case; it remains my responsibility until they’ve been seen.
>
> FG2: I think we’re all incredibly defensive. But that’s also partly how care is today, that we’re quite defensive. If you don’t, you’ll get into trouble because, of course, ‘the doctor should have known’.

## Discussion

The aim of this study was, first, to identify the healthcare needs referred to the internal medicine department that were redirected to the GP after triage, and, second, to identify the drivers underlying a GP referral.

In this study, the highest rate of referrals deemed unnecessary by triaging specialists was seen in the internal medicine domain endocrinology (35%). Geriatrics, ‘other’ and nephrology were also domains with high redirection rates. These redirected referrals were resolved in various ways, often including advice and guidance for the GP. It is important to consider the period this study was performed, the second year of triaging was in the middle of the COVID-19 pandemic. Referrals to hospitals may have been influenced, but also the triaging decisions by medical specialists could be influenced. It was tried to incorporate this by adding another six months in 2021. But even though referral behaviour may have been different, still 17% was deemed not necessarily secondary care. Qualitative results revealed two overarching domains influencing a GP’s decision to refer: 1) medical and 2) non-medical. These domains encompassed four themes, with the medical domain consisting of exclusively medical drivers (Theme 1), while the non-medical domain covered general practitioner-related drivers (Theme 2), patient-related drivers (Theme 3) and external drivers (Theme 4).

### Referring to internal medicine

Internal medicine is a diverse and multifaceted specialty, and sometimes the ‘last resort’ for referral (12). When GPs cannot classify a patient’s issue under a specific specialty, they often select ‘other’, contributing to a high rate of such referrals. However, referral to internal medicine also occurs due to multimorbidity, when patients do not fit a single category (13–17). This is similar to the 20% of “other” we found. Two-thirds of endocrinology referrals concerned thyroid issues or diabetes mellitus. Our focus groups revealed that, especially for diabetes, treatment options and guidelines are changing and developing rapidly (18). New drugs and new indications are continually becoming available, which makes treatment more complex. GPs reported difficulty keeping up with the pace of new developments and the changes in treatment responsibilities included in guidelines, which was also found in other research (19). Most redirected geriatric referrals concerned mild cognitive impairment or memory problems, which GPs are generally able and permitted to diagnose themselves, or alternatively consult an elderly care physician. As concerns nephrology, participating GPs stated that they always find the subject challenging, and despite the guideline tables presenting different treatment options, they still find it hard to manage or change treatments, as is shown in other studies that adhering to guidelines is not always the case (20).

Although triaging specialists judged a substantial proportion of these referrals to be unnecessary for secondary care, many still contained specific clinical questions or patient-related concerns that required clarification. For this reason, redirected referrals were often accompanied by targeted advice and guidance for the GP, rather than being returned without further support. This reflects an emerging interface model in which specialist input is provided without the need for face-to-face consultation. At the start of the study, video consultations and structured electronic consultations were not yet common practice; however, these modalities expanded rapidly during the COVID-19 pandemic. In early 2021, triaging specialists advised GPs to use an electronic consultation instead of submitting a standard referral, illustrating the developing role of digital tools in managing such cases.

### Drivers influencing the decision for referral

In line with our findings, earlier studies identified influences of both medical and non-medical factors on referral decisions in general practice (21–24). Notably, in both our and earlier studies, non-medical drivers often outweighed purely clinical indications. These included patient expectations, social context, as well as perceived relational dynamics. In particular, some studies found that referral decisions were frequently shaped by intuitive judgments and affective responses rather than structured clinical reasoning (21, 24). This corresponds with our observation that some reasons for referral could not be precisely defined or categorized, but rather seemed to be a ‘gut feeling’ or emotional undercurrent rather than an objective clinical necessity. Other research further elaborated on how individual interpretation and personal decision-making styles play a crucial role in whether and when GPs refer, reinforcing the subjective and situational nature of referral behaviour (23).

#### Medical domain Medical drivers

The most important medical reasons for referring were following or complying with Dutch GP guidelines or being in need of secondary specialist care. Results from another study, however, showed that almost 10% of redirected referrals could be attributed to ‘not complying with guidelines’ (25), and when a GP deviated from guidelines it was usually driven by a non-medical factor (26). GPs also rely on gut feeling and when alarmed they usually act (27–33), for example by ordering additional tests or referring to secondary care. Focus group participants stated that a referral is almost always accepted in cases where a GP consults a medical specialist based on this sense of alarm.

#### Non-medical domain

##### General practitioner-related drivers

Several studies have explored GP-related factors influencing referral decisions (21, 34–38). Based on our focus group results, we distinguished between *being* an autonomous healthcare professional, including the emotional dimension of doctor-patient relationships and *acting* as a healthcare provider who, for instance, experiences uncertainty about a treatment decision. A similar distinction was identified in another study, which categorised referral decisions as driven by either confidence or uncertainty (39). In our own study, uncertainty also emerged as a strong driver. This usually overlapped with other non-medical drivers, but also with medical drivers (3, 21, 22, 40). The presence or absence of confidence appears closely linked to variability in clinical reasoning, and in turn, referral patterns (24). Comparable to our findings, another study also found that uncertainty sometimes results in immediate referral whilst others prefer to ‘wait and see’ (21). This uncertainty was linked to (insufficient) experience and knowledge (gaps). For example, younger or less experienced GPs tended to refer earlier than the more experienced GPs. Endocrine referrals in our study were often attributable to uncertainty regarding current treatment or laboratory results. Another prominent driver was the importance of maintaining a good doctor–patient relationship. The trust and sense of responsibility embedded in this relationship were frequently cited as reasons for or against referral. GPs were often careful to preserve this trust, as maintaining it was seen as central to effective care (37, 41). However, this concern sometimes conflicted with their sense of professional autonomy.

#### Patient-related drivers

Almost all participating GPs noted terms like *‘patient pressure’*, *‘patient demand’*, or *‘patient wish’* on their sticky notes. All could immediately recall instances where patients had requested or even insisted on being referred to secondary care. Feeling pressured to refer is not new; it was described in studies over 30 years ago and remains relevant (21, 34, 42–47). Our participating GPs recognised this pressure from their own practice, although it varied across patients and situations. Sometimes they gave in to such pressure, but many actively tried to avoid it especially when they believed the referral was unnecessary (42, 48). As mentioned above, while doctor–patient relationships are important, clinicians rated their relationships as poorer in cases of referral following pressure (45). However, perceptions of patient pressure are subjective and appear to differ between GPs and to depend on the specialty being requested (49). Patient demographics were rarely mentioned in our focus groups, aside from the issue of language barriers in one session.

#### External drivers

Over the past decade, healthcare reforms have shifted care from secondary to primary care settings, aiming to reduce need for more expensive forms of treatment (11, 50, 51). Despite mixed results, interventions and pilot programs continue to promote this transition of care (52–56). GPs in our study reported experiencing this shift, noting increased responsibility within the same limited consultation times, and the need for broader medical knowledge across various medical disciplines. This sometimes led to an increased readiness to refer. This shift in care also contributes to a growing sense of time pressure and an inability to consistently provide thorough and accurate care. Alongside increased workload, new primary care models introduced changes in responsibility and legal obligations. Healthcare insurers and regulatory requirements have introduced new referral requirements (57). In the Netherlands, GPs remain responsible for patients even after a referral has been made, a responsibility that continues until the patient has had their first outpatient consultation (58). This situation frustrated several participants in our study, and they emphasized that a referral is a clear signal of their need for specialist support. However, due to long waiting lists they often remain responsible for patients they can no longer adequately help.

With the expected rise in the ageing population, multimorbidity and chronic diseases, combined with a reduced workforce, it is vital to provide care at the most appropriate time and place (11). Collaboration between GPs and specialists is an important factor in achieving this. Successful collaboration requires clarity regarding which patients need face-to-face specialist care and which ones can be appropriately managed within general practice, possibly supported by advice from secondary care. Preventing unnecessary care can by itself be considered an improvement in patient care, as it allows patients to continue treatment with their trusted primary care provider. For urgent or critical cases, fast-track outpatient services enable prompt assessment and management, addressing potential risks associated with waiting times. Furthermore, managing increasingly complex diseases in primary care is becoming more challenging. The decision-making process underlying a GP’s choice to refer remains a complex and difficult-to-define phenomenon. Various factors interact continuously, and each can vary across time, context and individual GP.

Future research should focus on further optimising the interface between primary and secondary care, including the use of electronic and video consultations to support GPs in managing referrals. Evaluating the impact of such initiatives on patient outcomes, satisfaction, accessibility, and safety will be essential. Additionally, exploring the scalability and implementation of this approach across different regions and medical specialties could provide valuable insights for broader application. Generalisability to other countries should be researched because it might be limited and partly acceptable in countries with similar GP and primary- secondary care systems. However, this study also provides broad insights into organising referral management.

## Conclusion

This study provides a deeper understanding of the needs of primary care referrals to internal medicine. To optimize triaging of GP referrals to secondary care, it is essential to understand the specific motivations underlying each referral. What became evident during this study is the value of deeper insights into the reasons underlying GP referrals, especially the interplay of both medical and non-medical factors in clinical reasoning. Nonetheless, our finding that 17% of internal medicine referrals could potentially be managed in primary care is remarkable and highlights an opportunity to reconsider and reshape the interface between GPs and specialists. Insight into this reasoning will support mutual understanding and collaboration, helping to ensure and safeguard high-quality patient care in both primary and secondary care.

## Data Availability

All data produced in the present study are available upon reasonable request to the authors

## Acknowledgements

We would like to thank all general practitioners who participated in the study for sharing their honest opinions and experiences. Thanks to them, this manuscript now contains in-depth information about referrals and primary practice. We would also like to express our sincere gratitude to Chakir Mousaoui for his help and dedication in organising the focus groups, as well as his assistance with the coding and analysis of the transcripts.

